## Supplementary information for "Transmission dynamics of *Klebsiella pneumoniae* in a neonatal intensive care unit in Zambia before and after an infection control bundle"

### Table S1: Reference sequence accessions for each ST

| Sequence type | Reference accession |
|---|---|
| ST15 | NZ_CP062475.1 |
| ST101 | NZ_CP102940.1 |
| ST147 | NZ_CP029582.1 |
| ST307 | NZ_CP158302.1 |
| ST983 | NZ_CP021165.1 |
| ST985 | NZ_CP086724.1 |
| ST2004 | NZ_CP103579.1 |

### Table S2: Isolates and sequence data included in the study

https://github.com/klebgenomics/SPINZ/blob/main/tables/Supplementary_table_S2.tsv

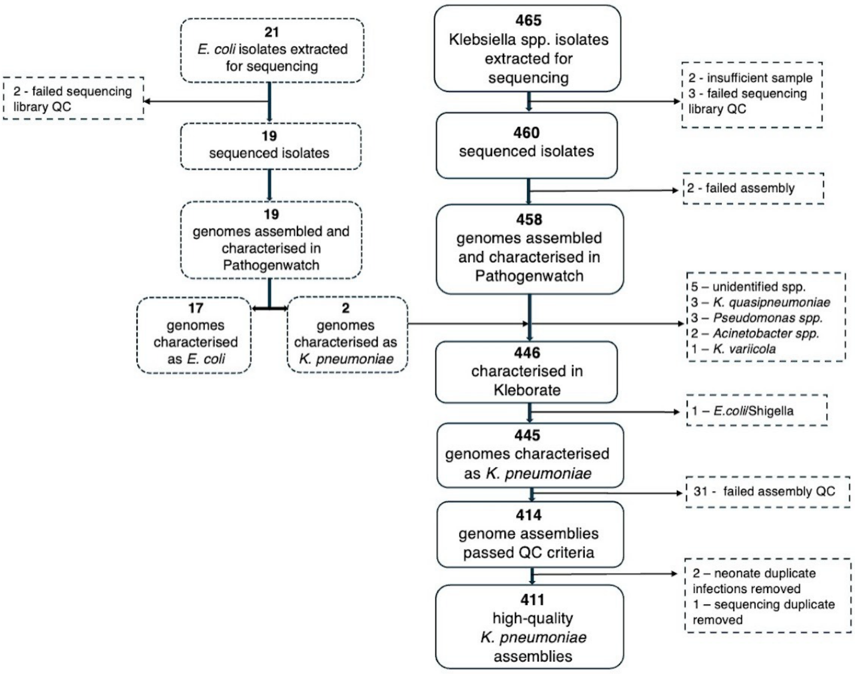


**Figure S1. Pipeline of isolate from isolate extraction through to high-quality assembled genomes.** Loss of isolates from the pipeline are indicated at each stage.

| ST307 | | Temporal threshold (weeks) | | | |
|---|---|---|---|---|---|
| | | **1** | **2** | **3** | **4** |
| **SNP distance threshold 5-20** | **No of clusters** | 8 | 7 | 5 | 5 |
| | **No of clustered sequences** | 225/232 | 227/232 | 230/232 | 231/232 |
| | **Median cluster size** | 7 | 7 | 7 | 7 |
| | **Proportion of cases in a transmission cluster** | 97% | 98% | 99% | 100% |
| | **Proportion of cases due to transmission** | 94% | 95% | 97% | 97% |

| ST101 | | Temporal threshold (weeks) | | | |
|---|---|---|---|---|---|
| | | **1** | **2** | **3** | **4** |
| **SNP distance threshold 5** | **No of clusters** | 1 | 2 | 2 | 2 |
| | **No of clustered sequences** | 2/9 | 4/9 | 4/9 | 5/9 |
| | **Median cluster size** | 2 | 2 | 2 | 2 |
| | **Proportion of cases in a transmission cluster** | 22% | 44% | 44% | 56% |
| | **Proportion of cases due to transmission** | 11% | 22% | 22% | 33% |
| **SNP distance threshold 10 - 25** | **No of clusters** | 1 | 2 | 2 | 2 |
| | **No of clustered sequences** | 2/9 | 5/9 | 6/9 | 6/9 |
| | **Median cluster size** | 2 | 2 | 3 | 3 |
| | **Proportion of cases in a transmission cluster** | 22% | 56% | 67% | 67% |
| | **Proportion of cases due to transmission** | 11% | 33% | 44% | 44% |

| ST2004 | | Temporal threshold (weeks) | | | |
|---|---|---|---|---|---|
| | | **1** | **2** | **3** | **4** |
| **SNP distance threshold 5-25** | **No of clusters** | 6 | 2 | 2 | 2 |
| | **No of clustered sequences** | 44/47 | 47/47 | 47/47 | 47/47 |
| | **Median cluster size** | 4 | 24 | 24 | 24 |
| | **Proportion of cases in a transmission cluster** | 94% | 100% | 100% | 100% |
| | **Proportion of cases due to transmission** | 81% | 96% | 96% | 96% |

| ST983 | | Temporal threshold (weeks) | | | |
|---|---|---|---|---|---|
| | | **1** | **2** | **3** | **4** |
| **SNP distance threshold 5-25** | **No of clusters** | 2 | 1 | 1 | 1 |
| | **No of clustered sequences** | 9/9 | 9/9 | 9/9 | 9/9 |
| | **Median cluster size** | 4 | 9 | 9 | 9 |
| | **Proportion of cases in a transmission cluster** | 100% | 100% | 100% | 100% |
| | **Proportion of cases due to transmission** | 78% | 89% | 89% | 89% |

| ST147 | | Temporal threshold (weeks) | | | |
|---|---|---|---|---|---|
| | | **1** | **2** | **3** | **4** |
| **SNP distance threshold 5-25** | **No of clusters** | 1 | 1 | 1 | 1 |
| | **No of clustered sequences** | 3/4 | 3/4 | 4/4 | 4/4 |
| | **Median cluster size** | 3 | 3 | 4 | 4 |
| | **Proportion of cases in a transmission cluster** | 75% | 75% | 100% | 100% |
| | **Proportion of cases due to transmission** | 50% | 50% | 75% | 75% |

| | ST15 | **Temporal threshold (weeks)** | ST985 | **Temporal threshold (weeks)** |
|---|---|---|---|---|
| | | **1-4** | | **1-4** |
| **SNP distance threshold 5-25** | **No of clusters** | 1 | **No of clusters** | 1 |
| | **No of clustered sequences** | 4/4 | **No of clustered sequences** | 5/5 |
| | **Median cluster size** | 4 | **Median cluster size** | 5 |
| | **Proportion of cases in a transmission cluster** | 100% | **Proportion of cases in a transmission cluster** | 100% |
| | **Proportion of cases due to transmission** | 75% | **Proportion of cases due to transmission** | 80% |

| ST985 | | **Temporal threshold (weeks)** |
| --- | --- | --- |
| | | **1-4** |
| **SNP distance threshold 5-25** | **No of clusters** | 1 |
| | **No of clustered sequences** | 5/5 |
| | **Median cluster size** | 5 |
| | **Proportion of cases in a transmission cluster** | 100 |
| | **Proportion of cases due to transmission** | 80 |

**Figure S2. Clustering sensitivity analyses.** The sensitivity of clustering to choice of SNV threshold (between 5–25) and temporal threshold (between 7–28 days) was assessed for all STs.

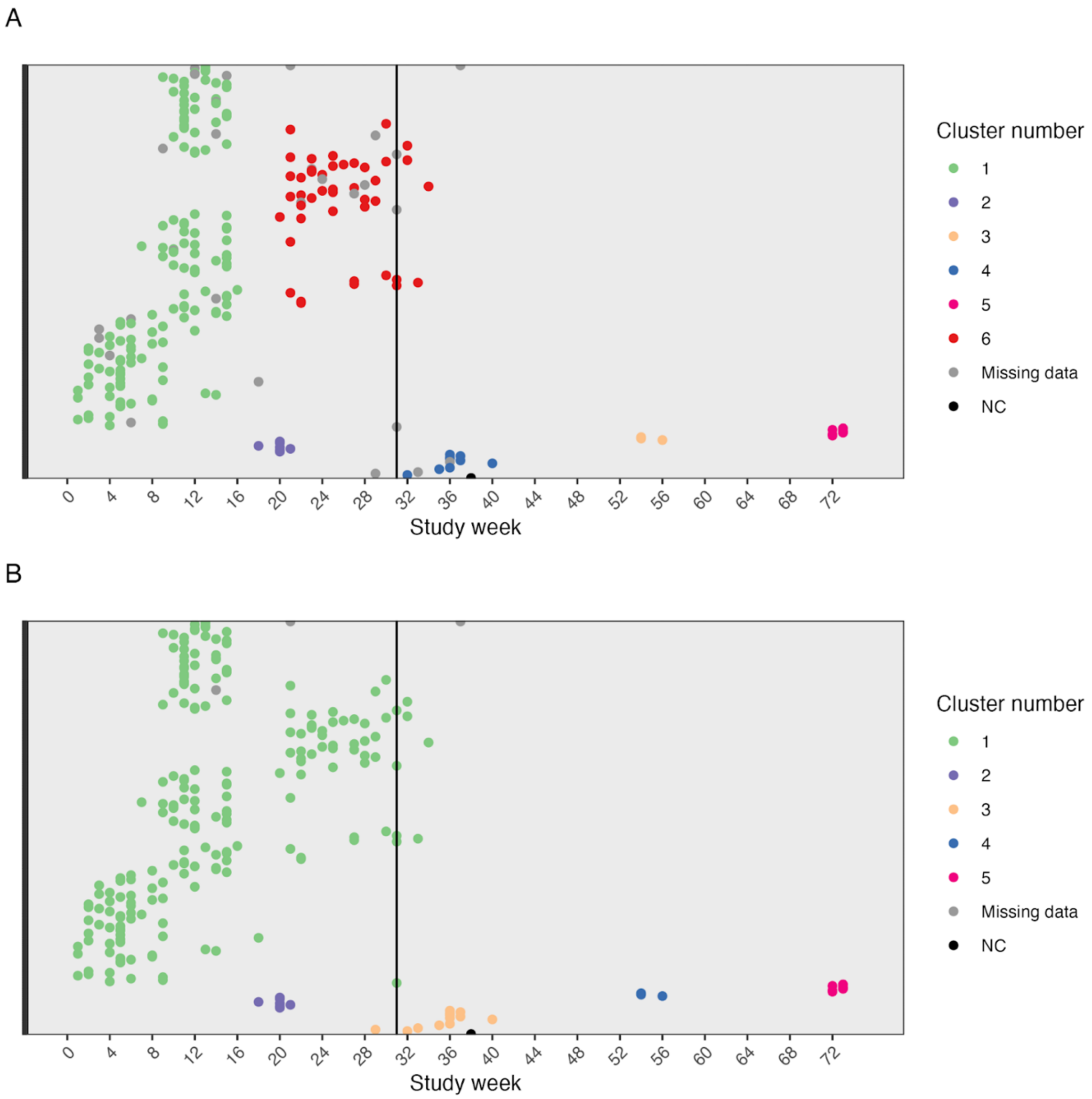


**Figure S3. Sensitivity analysis of ST307 clusters. (A)** Using culture date to define clusters if available, and admission date if not (n=233), compared to **(B)** using only culture date (n=206).

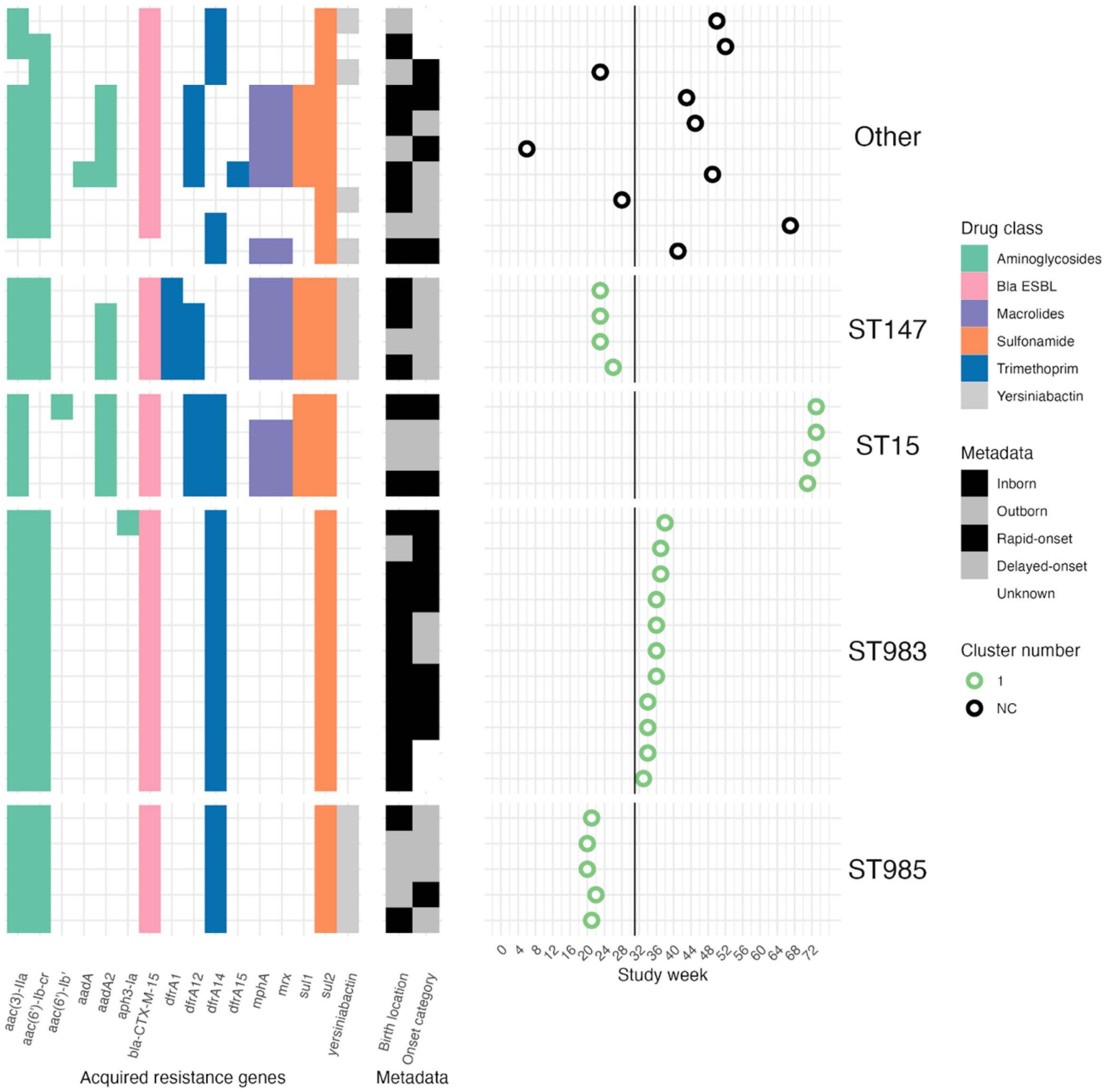


**Figure S4. Acquired resistance genes, hypervirulence genes and birth location and onset metadata aligned to study week of infection for all other STs (of ≥4 genomes).** In all other STs for which a phylogenetic tree could not be constructed. The presence of an isolate in a cluster is indicated by the colour of the circle plotted by study week of infection. The solid vertical line represents the start of the implementation of the IPC bundle.

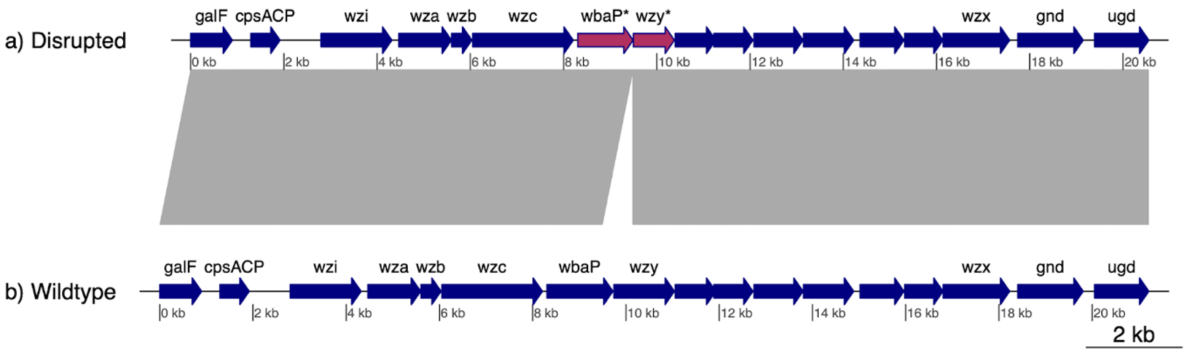

**Figure S5. Comparison of KL102 loci in ST307. (a)** Disrupted form conserved in ST307 cluster 1 isolates from this study. **(b)** Wildtype reference form (GenBank accession, AB371290). Arrows show annotated protein-coding genes, navy blue arrows indicate intact open reading frames, maroon arrows indicate truncated open reading frames. Grey shows homology blocks between the two KL102 locus sequences, revealing a 661 bp deletion stretching from the end of wbaP into the beginning of wzy thus truncating both genes and presumably rendering both non-functional.

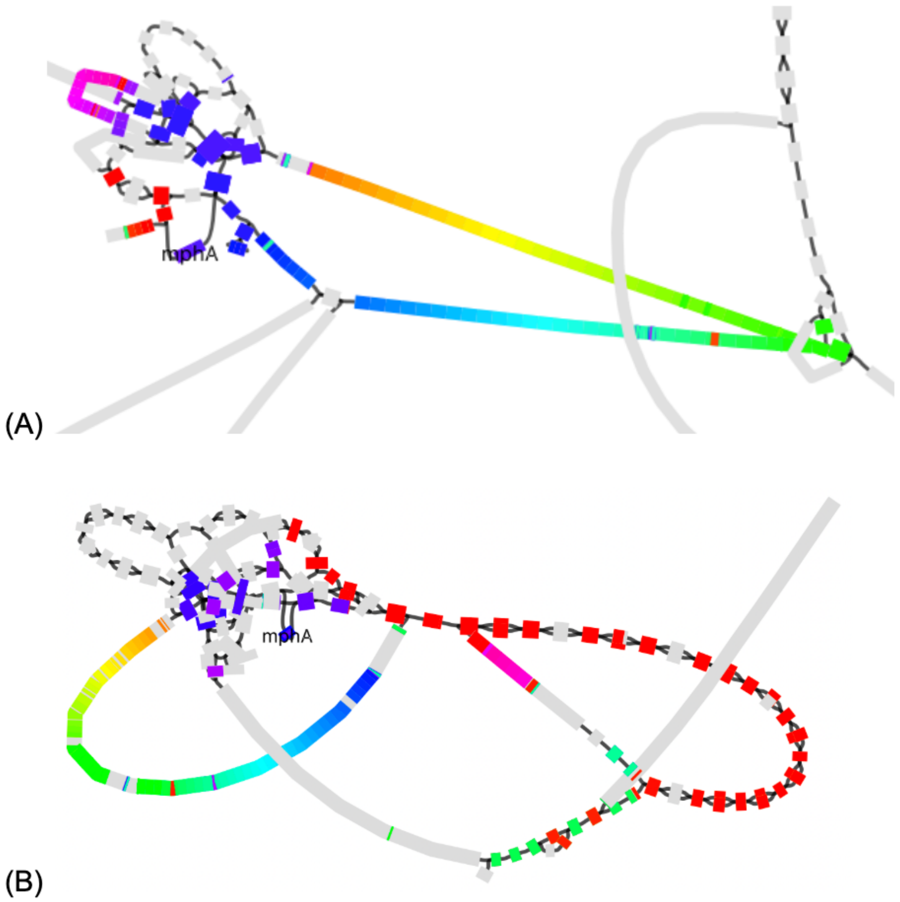


**Figure S6. BLAST hits to mphA reference plasmid (GenBank accession: CP021752.1) in representative assembly graphs for ST307 cluster 3 and 4 genomes.** BLAST search and visualisation of the graph was done using Bandage. In each graph, rectangles indicate contigs, sized to indicate their relative length in base pairs. Contigs are coloured to indicate hits to the reference plasmid (GenBank accession CP021752.1), using a rainbow scale such that red indicates hits to the start of the reference plasmid sequence, colours yellow through green indicate hits in the middle of the reference sequence, through to blue for hits at the end of the reference sequence. The contig containing the mphA gene is labelled. (A) Cluster 4 genome, read accession ERR15165518. Zoomed in on the part of the graph with hits to the reference plasmid (96.9% coverage at mean 98.6% identity), note this part is connected to the rest of the graph including the chromosome. (B) Cluster 3 genome, ZN2356 (accession ERR15165559). Note the part of the graph shown is disconnected from the rest of the graph including the chromosome. Coverage of the reference plasmid is 74%.

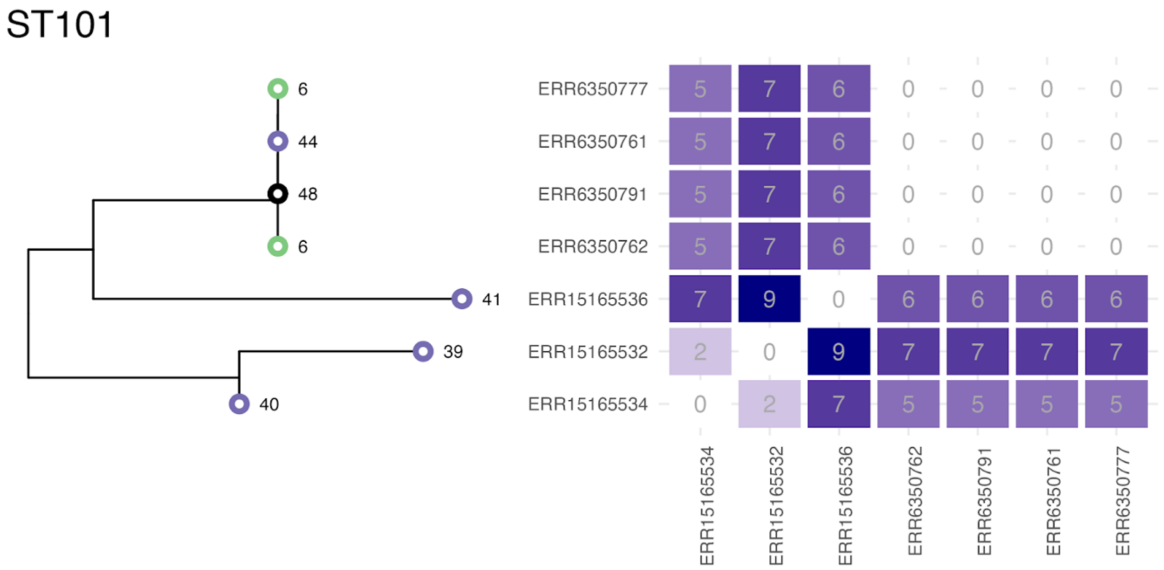


**Figure S7. Maximum likelihood phylogenetic tree and pairwise SNV distance matrix of a subset of ST101 infections.** Tree tips labelled with cluster number (colour) and week of infection. Two infections observed in week 6 during the baseline period (cluster 1, green tree tips), and four infections during weeks 39-44 in the post-implementation period (cluster 2, purple tree tips) as well as a single non-clustered infection (black tree tip) at week 48.
